# High-Resolution Mapping of Healthcare Pathways in Autism

**DOI:** 10.64898/2026.10.30.26366679

**Authors:** Like Huang, Tai Ren, Can Hou, Fang Fang, Jakob Grove, Zhenzhen Xie, Yiwei Pu, Diana Schendel, Brian K. Lee, Fei Li, Viktor H. Ahlqvist

## Abstract

Autistic children and adolescents experience substantial psychiatric and somatic comorbidity, but whether this excess morbidity follows structured diagnostic patterns or temporal trajectories remains unclear. We conducted a nationwide register-based study of 101,521 autistic individuals in Sweden and 958,093 age- and sex-matched peers, followed from birth through early life until a median age of 19.8 years. Across 12.86 million diagnostic codes from 9.49 million inpatient and specialist outpatient healthcare encounters, autistic individuals had increased risk of 165 of 187 examined conditions. Excess morbidity was not diffusely distributed across the phenome, but clustered into seven distinct, system-dominant domains. Temporal trajectory analyses further showed that autism-enriched diagnostic sequences were concentrated within respiratory, congenital, and mental–injury/poisoning-related domains. These findings indicate that morbidity in autistic young people is both clinically structured and temporally organized, providing a population-level framework for understanding the concentrated domains that drive excess healthcare burden.

## Introduction

Individuals with autism face elevated risks of both psychiatric and somatic comorbidities, contributing to an excess premature mortality(1–4). Children and young people with autism, in particular, are frequently diagnosed with psychiatric disorders, co-occurring neurological conditions, and somatic diseases(5–7). In Sweden, these comorbidities may contribute to the 16.3-year lower life expectancy observed in individuals with autism(8).

Yet the existing evidence base has largely examined these conditions in isolation(1, 9–12), focusing on single diagnoses as outcomes rather than the interconnected pathways that shape real-world health and healthcare. In practice, comorbidities are seldom isolated, and care trajectories rarely follow a linear course. For autistic individuals, navigating healthcare systems may be further complicated by sensory sensitivities, communication differences, executive functioning challenges, stigma, and insufficient provider training (13–15).

To advance our understanding of pathways of co-occurring health conditions, researchers have begun moving beyond condition-by-condition analyses to capture how comorbidities cluster and evolve over time(16–18). Recent advances in availability of large-scale electronic health records data, high-performance computing, and machine learning enable such analyses, providing opportunities to map patient flows, identify common pathways, and reveal previously unrecognized patterns of comorbidity co-occurrence (17). These approaches may refine our knowledge of established comorbidities, such as psychiatric, neurological, and cardiovascular diseases in autistic individuals, but also generate novel insights into the dynamic health trajectories that individuals with autism experience.

Here, we leverage our recently developed analytic framework that integrates phenome-wide association (PheWAS), comorbidity network modelling, and disease trajectory analysis(19, 20), applied to Sweden’s comprehensive national healthcare registers. Leveraging the rich data with millions of healthcare interactions, this approach moves beyond isolated associations to generate a multidimensional representation of autistic individuals’ healthcare experiences. Our goal is to deliver a high-resolution characterization of healthcare pathways in autism - laying the groundwork for a deeper understanding of needs and for the development of interventions that can anticipate, prevent, and, ultimately, mitigate adverse outcomes.

## Results

We constructed a nationwide matched cohort comprising all individuals born in Sweden between 1987 and 2020 with a diagnosis of autism any time through 31 December 2021 (N = 101,521). Each individual diagnosed with autism was age and sex matched with up to 10 unaffected individuals. These unaffected peers were required to be alive and residing in Sweden on the diagnosis date of the corresponding ‘case’ and to have no recorded autism diagnosis throughout follow-up (N = 958,093). We then proceeded to follow this cohort throughout Swedish healthcare records (inpatient and specialized outpatient) (Figure 1), following everyone from birth until death, emigration, or the end of follow-up (31 December 2021), which-ever occurred first.

**Figure 1.**
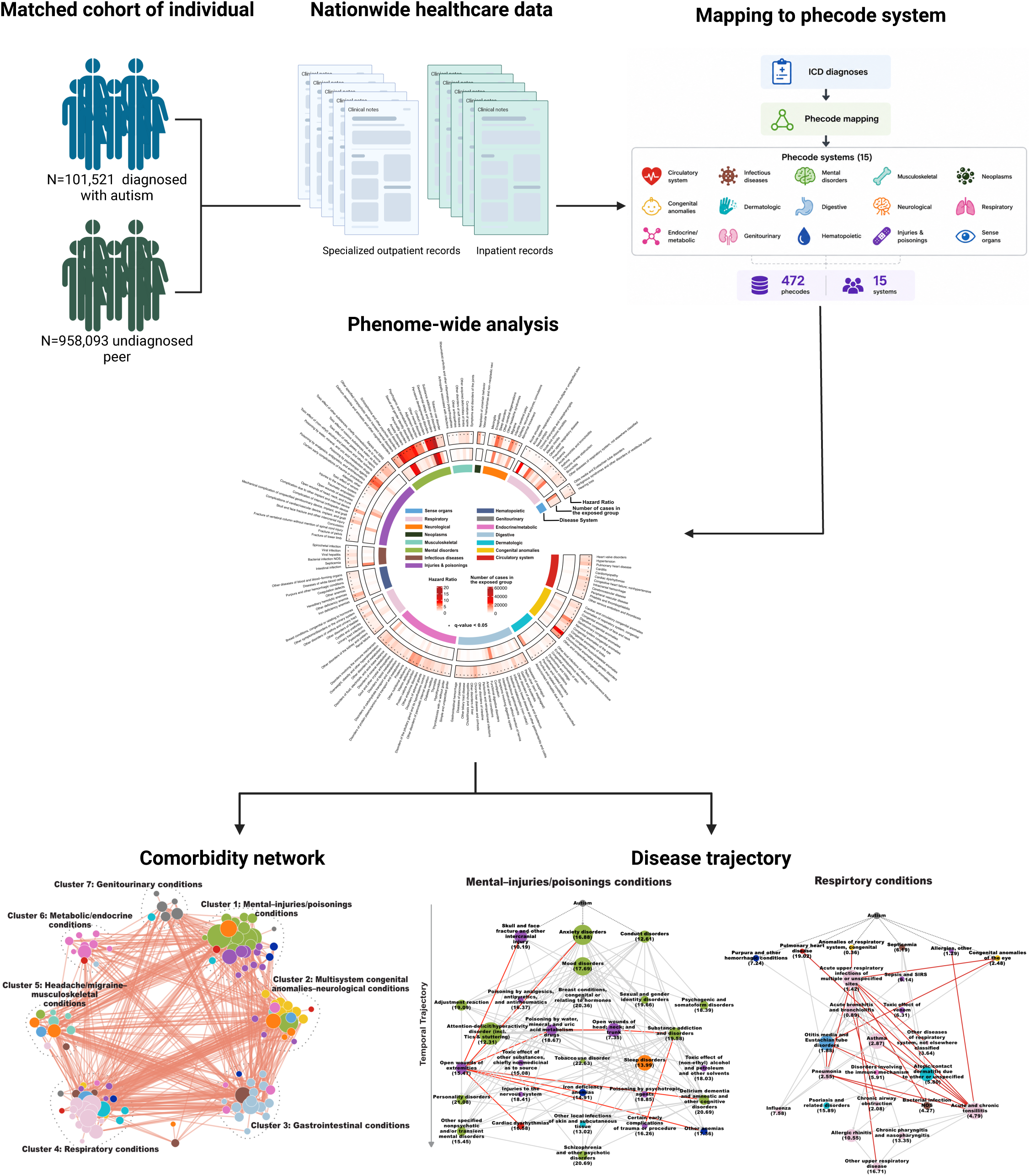
Overview of the study design and analytic pipeline. Overview of the study design and analytic pipeline. The study was based on a nationwide matched cohort comprising 101,521 individuals diagnosed with autism and 958,093 matched unaffected peers. Nationwide healthcare data, including specialized outpatient and inpatient records, were used to identify medical conditions. Diagnostic codes were mapped to the phecode system and analyzed using phenome-wide analysis, comorbidity network analysis, and disease trajectory analysis to characterize co-occurring conditions and their patterns.

Among the 1,059,614 included individuals (Table 1), those diagnosed with autism were more likely to be born in Sweden (93.0% vs. 79.9%) and to have complete data of parental education and family income, as compared with the unaffected peers. The follow-up periods were comparable between the two groups (median, 19.9 years [interquartile range 14.4–26.5] for individuals diagnosed with autism, 19.8 years [interquartile range 14.2–26.3] for the unaffected peers).

**Table 1.** Baseline characteristics of study participants.

| Characteristics | Individuals diagnosed<br>with autism<br>(N=101,521) | Matched individuals<br>with no diagnosis of au-<br>tism<br>(N=958,093) |
| --- | --- | --- |
| **Sex, n (%)** |  |  |
| Male | 66,606 (65.61%) | 630,696 (65.83%) |
| Female | 34,915 (34.39%) | 327,397 (34.17%) |
| **Birth year, n (%)** |  |  |
| 1987-1990 | 10,561 (10.40%) | 96,681 (10.09%) |
| 1991-1995 | 17,209 (16.95%) | 160,659 (16.77%) |
| 1996-2000 | 18,603 (18.32%) | 175,722 (18.34%) |
| 2001-2005 | 23,060 (22.71%) | 217,552 (22.71%) |
| 2006-2010 | 18,720 (18.44%) | 177,896 (18.57%) |
| 2011-2015 | 10,536 (10.38%) | 101,797 (10.62%) |
| 2016-2020 | 2,832 (2.79%) | 27,786 (2.90%) |
| **Country of birth, n (%)** |  |  |
| Sweden | 94,452 (93.04%) | 765,398 (79.89%) |
| Outside Sweden | 7,069 (6.96%) | 192,695 (20.11%) |
| **Highest parental attained education, n (%) <sup>a</sup>** |  |  |
| 1 (lowest) | 5,612 (5.53%) | 51,630 (5.39%) |
| 2 | 7,074 (6.97%) | 61,464 (6.42%) |
| 3 | 15,267 (15.04%) | 136,043 (14.20%) |
| 4 | 20,835 (20.52%) | 179,143 (18.70%) |
| 5 | 15,677 (15.44%) | 136,610 (14.26%) |
| 6 | 28,020 (27.60%) | 241,232 (25.18%) |
| 7 (highest) | 1,796 (1.77%) | 15,404 (1.61%) |
| Missing | 7,240 (7.13%) | 136,567 (14.25%) |
| **Family income quintile (n, %) <sup>b</sup>** |  |  |
| 1 (lowest) | 19,258 (18.97%) | 167,486 (17.48%) |
| 2 | 19,068 (18.78%) | 167,734 (17.51%) |
| 3 | 19,253 (18.96%) | 168,036 (17.54%) |
| 4 | 19,337 (19.05%) | 167,931 (17.53%) |
| 5 (highest) | 19,410 (19.12%) | 168,006 (17.54%) |
| Missing | 5,195 (5.12%) | 118,900 (12.41%) |
| **Follow-up time in years (median, IQR)** | 19.88 (14.34-26.54) | 19.76 (14.20-26.26) |

### Phenome-wide association results

During the follow-up, the cohort had 9.49 million inpatient and specialist outpatient interactions, spanning 12.86 million diagnostic codes recorded using International Classification of Diseases, Ninth Revision (ICD-9) or Tenth Revision (ICD-10) codes. All codes were mapped into 472 clinically meaningful phenotypes (“phecodes”, hereinafter also referred to as medical conditions), according to a standardized catalog developed for PheWAS in large electronic health record datasets(21, 22). For each medical condition, incident co-occurring morbidity was defined as the first recorded diagnosis in inpatient or outpatient specialist care.

Among the 187 medical conditions with an incidence of ≥0.1% among individuals diagnosed with autism (a criterion implemented to filter away rare conditions), 165 were more prevalent among autistic individuals compared to the matched unaffected peers (Benjamini-Hochberg (BH) adjusted P<0.05; Figure 2; Table S1). The condition with the highest cumulative incidence by age 20 was the ADHD phecode (which covers ADHD, tics, and stuttering; 90% of the cases under this phecode was ADHD), affecting 51.7% of the autism group compared to 6.5% of the unaffected peers. Other medical conditions with a high cumulative incidence by age 20 in the autism group included anxiety disorders (35.5% vs 5.7%), developmental delays and disorders (30.2% vs 2.3%), mood disorders (27.2% vs 3.9%), sleep disorders (16.1% vs 2.7%), asthma (17.1% vs 11.6%), constipation (15.7% vs 7.4%), intestinal infection (14.5% vs 9.4%), and epilepsy, recurrent seizures, or convulsions (11.0% vs 3.3%).

**Figure 2.**
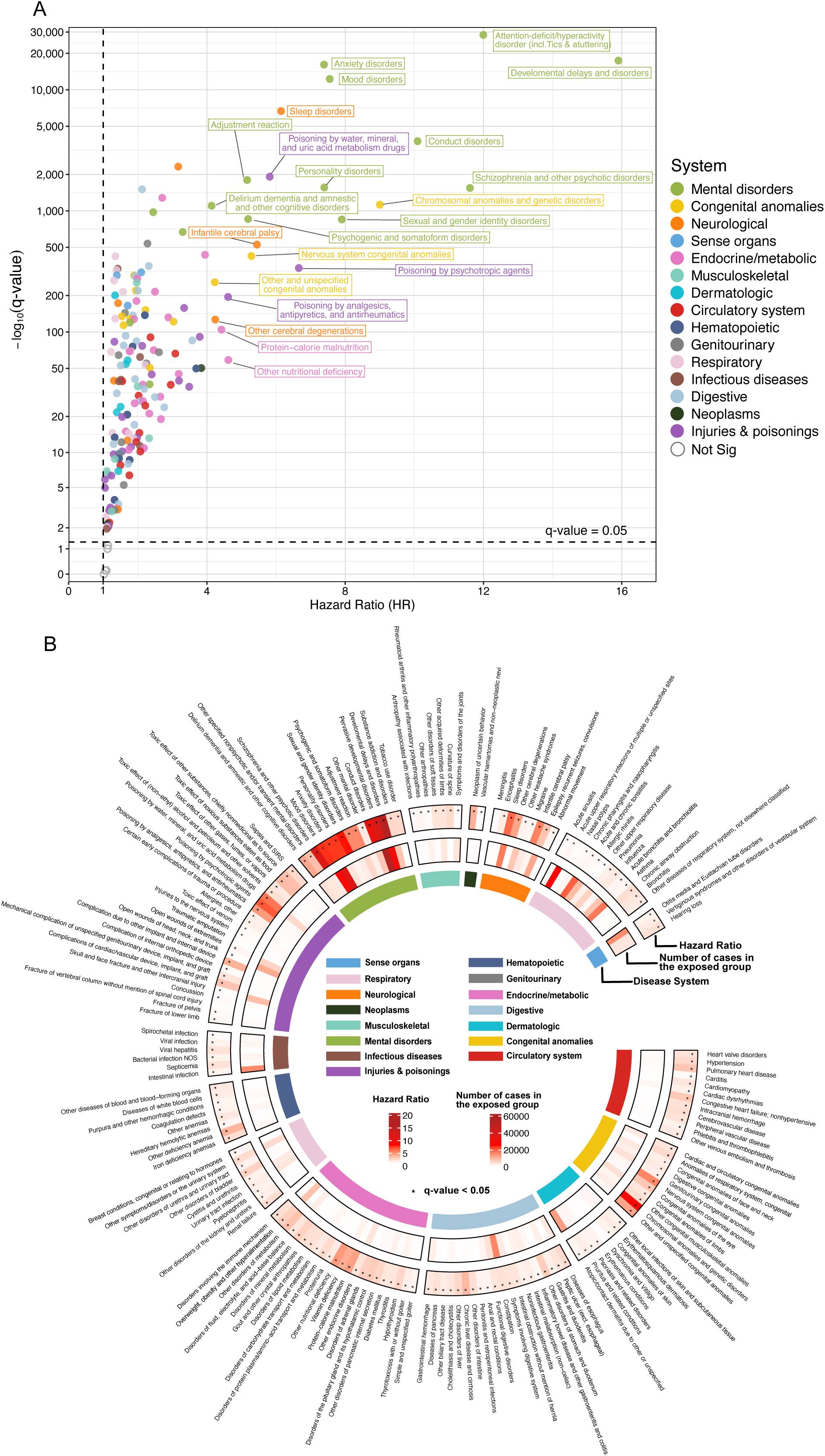
PheWAS analysis results for medical conditions comparing autistic individuals with matched autism-free individuals. (A) Scatter plot of hazard ratios (HRs) and corresponding significance levels for 187 phecode-defined medical conditions with a prevalence of ≥0.1% among autistic individuals and elevated risk in autism (HR >1). Each point represents one phecode-defined condition, with the x-axis indicating the HR and the y-axis showing the corresponding −log_10_(q value). Colors denote disease systems. The vertical dashed line indicates HR = 1, and the horizontal dashed line indicates the false discovery rate threshold of q = 0.05. Selected conditions with the strongest associations are labeled. (B) Circular summary plot of phenome-wide association results across disease systems. The inner colored ring denotes disease system categories, and each outer segment represents an individual medical condition. Color intensity indicates the magnitude of the HR, and the outer heatmap shows the number of cases in the autistic group. Asterisks indicate associations that remained significant after false discovery rate correction (q < 0.05).

After adjusting for matching variables (sex and birth year and month), country of birth, highest parental attained education and family income, the highest hazard ratios (HRs) were observed for mental disorders, including developmental delays and disorders (HR [95% CI]: 15.9 [15.6-16.2]), ADHD (including tics and stuttering) (12.0 [11.8-12.2]), schizophrenia and other psychotic disorders (11.6 [11.0-12.3]) and conduct disorders (10.1 [9.8-10.4]). High HRs were also noted for congenital anomalies, among which the strongest associations were noted for chromosomal anomalies and genetic disorders (9.0 [8.5-9.6]), and nervous system congenital anomalies (5.3 [4.9-5.7]). Neurological conditions with the highest HRs included sleep disorders (6.1 [6.0-6.3]), infantile cerebral palsy (5.4 [5.1-5.8]), other cerebral degenerations (4.2 [3.8-4.8]) and epilepsy, recurrent seizures, or convulsions (3.2 [3.1-3.2]). Poisonings-related conditions with the highest HRs included poisoning by psychotropic agents (6.7 [6.1-7.3]), poisoning by water, mineral, and uric acid metabolism drugs (5.8 [5.6-6.0]), and poisoning by analgesics, antipyretics, and antirheumatics (4.6 [4.2-5.1]). Elevated risks were also seen for endocrine/metabolic and nutrition-related conditions, including other nutritional deficiency (4.6 [3.8-5.6]), protein-calorie malnutrition (4.4 [3.8-5.1]), and vitamin deficiency (3.9 [3.7-4.2]).

### Comorbidity networks

To understand the clustering of comorbidities in autism we examined the associations between the 165 medical conditions with evidence of excess risk associated with autism. This led to identification of 2,079 disease pairs with some pairwise correlations within the group of individuals diagnosed with autism, out of the 13,530 possible pairwise combinations (i.e., 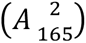). To eliminate pairs that were explained by some other comorbidity or covariates (birth country, highest parental education, family income, age at autism diagnosis, sex, or birth year), we performed Cox regression wherein we adjusted for the covariates and 20 principial components of derived from all other conditions than the two being examined. After this, 768 disease pairs out of the original 2,079 had statistically significant correlations (BH adjusted P<0.05) and were used to construct the comorbidity network (Figure 3A; Table S2).

**Figure 3.**
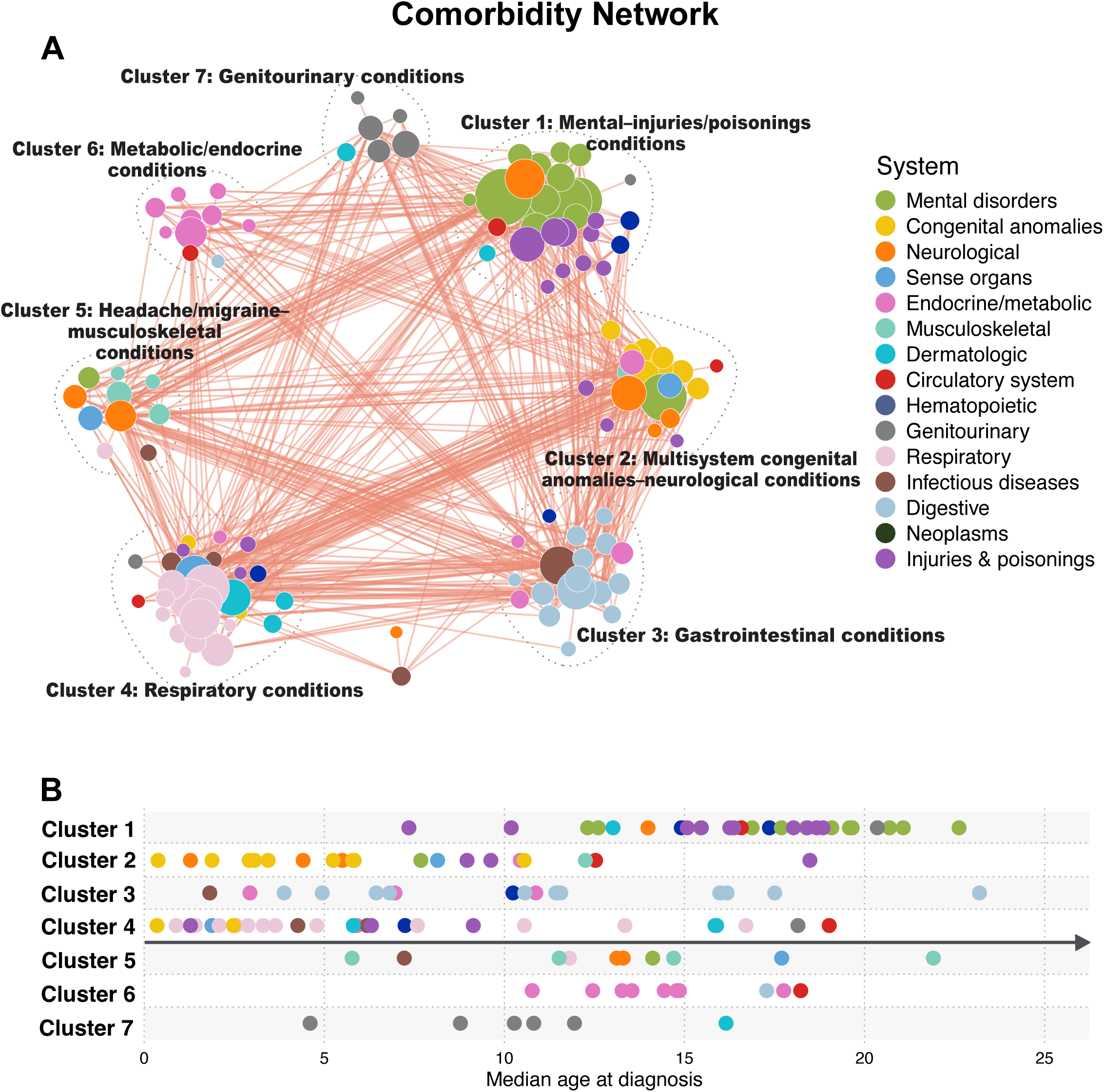
Comorbidity network of medical conditions among autistic individuals. (A) Comorbidity network of medical conditions among autistic individuals. Each node represents a phecode-defined medical condition, and node color indicates disease system. Edges represent significant co-occurrence relationships between conditions, and node size reflects the number of cases. (B) Distribution of the median age at diagnosis for conditions in each cluster. Each point represents one condition.

Using the Louvain algorithm on these 768 disease pairs we identified seven main clusters (Figure 3A; clusters were labeled post hoc according to their predominant condition types): i) a mental–injuries/poisonings cluster, ii) a multisystem congenital anomalies–neurological cluster, iii) a respiratory conditions cluster, iv) a gastrointestinal cluster, v) a headache/migraine–musculoskeletal cluster, vi) a metabolic/endocrine cluster, and vii) a genitourinary cluster. The median age of first diagnosis differed across clusters (Figure 3B). Conditions in the multisystem congenital anomalies–neurological (Figure S4B) and respiratory clusters (Figure S4D) were generally first diagnosed before 10 years of age. The median age of diagnosis was generally later than 10 years of age for conditions in the metabolic/endocrine (Figure S4F), mental–injuries/poisonings (Figure S4A), and headache/migraine–musculoskeletal clusters (Figure S4E).

To understand if the comorbidity networks were dominated by some condition that was correlated with everything else in the cluster, we computed the cluster-specific centrality of each condition (i.e., sum of edge weights that represented pairwise correlations). We found evidence of some conditions that showed correlations spanning multiple organ systems, such as intestinal infection, other headache syndromes, and pneumonia (Figure S3A).

Conditions within each of the seven clusters are largely onto shared organ systems. Developmental delay and epilepsy showed broad connectivity with multiple congenital anomalies (Figure S2B, 4B) and grouped within the multisystem congenital anomalies/neurological cluster rather than the mental–injuries/poisonings cluster (Figure S2A, 4A), underscoring their integration within early-onset neurodevelopmental pathology. The gastrointestinal cluster demonstrated the strongest cross-cluster connectivity, with the highest summed edge weights linking it to other clusters (Figure S3I). Injury and poisoning conditions co-clustered with mental–behavioral disorders (Figure S2A, 4A) and exhibited similar ages at first diagnosis (Figure S4A). Notably, two nutritional deficiencies (iron-deficiency anemia, and other anemias) also localized to this cluster rather than to gastrointestinal conditions (Figure S2A, 4A), potentially suggesting alignment with broader behavioral or systemic vulnerability patterns. Within the gastrointestinal cluster, intestinal infection demonstrated the highest centrality (Figures S3D, 4C), suggesting that it served as a key hub linking conditions within this cluster. The respiratory cluster was centered on pneumonia and atopic conditions, including asthma (Figures S3E, 4D). Although migraine is frequently reported to co-occur with psychiatric disorders, in this population it clustered preferentially with musculoskeletal conditions (Figures S3F, 4E). In the metabolic/endocrine cluster, diabetes mellitus exhibited the highest centrality (Figures S3G, 4F).

### Disease trajectories

To characterize the temporal emergence of comorbidity clusters, we next examine directional ordering between conditions. That is, across the 2,079 disease pairs, we assessed whether one condition was more likely to precede the other. In 469 pairs, one condition showed a significantly higher likelihood of preceding the other (BH adjusted P<0.05), after adjustment for country of birth, highest parental education, family income, age at autism diagnosis, sex, birth year, and the first 20 principal components derived from all other conditions.

Within the seven cross-sectionally defined clusters, 107 conditions could be temporally ordered (Figure 4; Table S3). For example, in the gastrointestinal cluster, early gastrointestinal infections were commonly followed by constipation, with subsequent progression to chronic inflammatory and functional conditions in adolescence. The metabolic–endocrine cluster was anchored by overweight and obesity as central nodes, with prominent temporal links to a range of cardiometabolic conditions.

**Figure 4.**
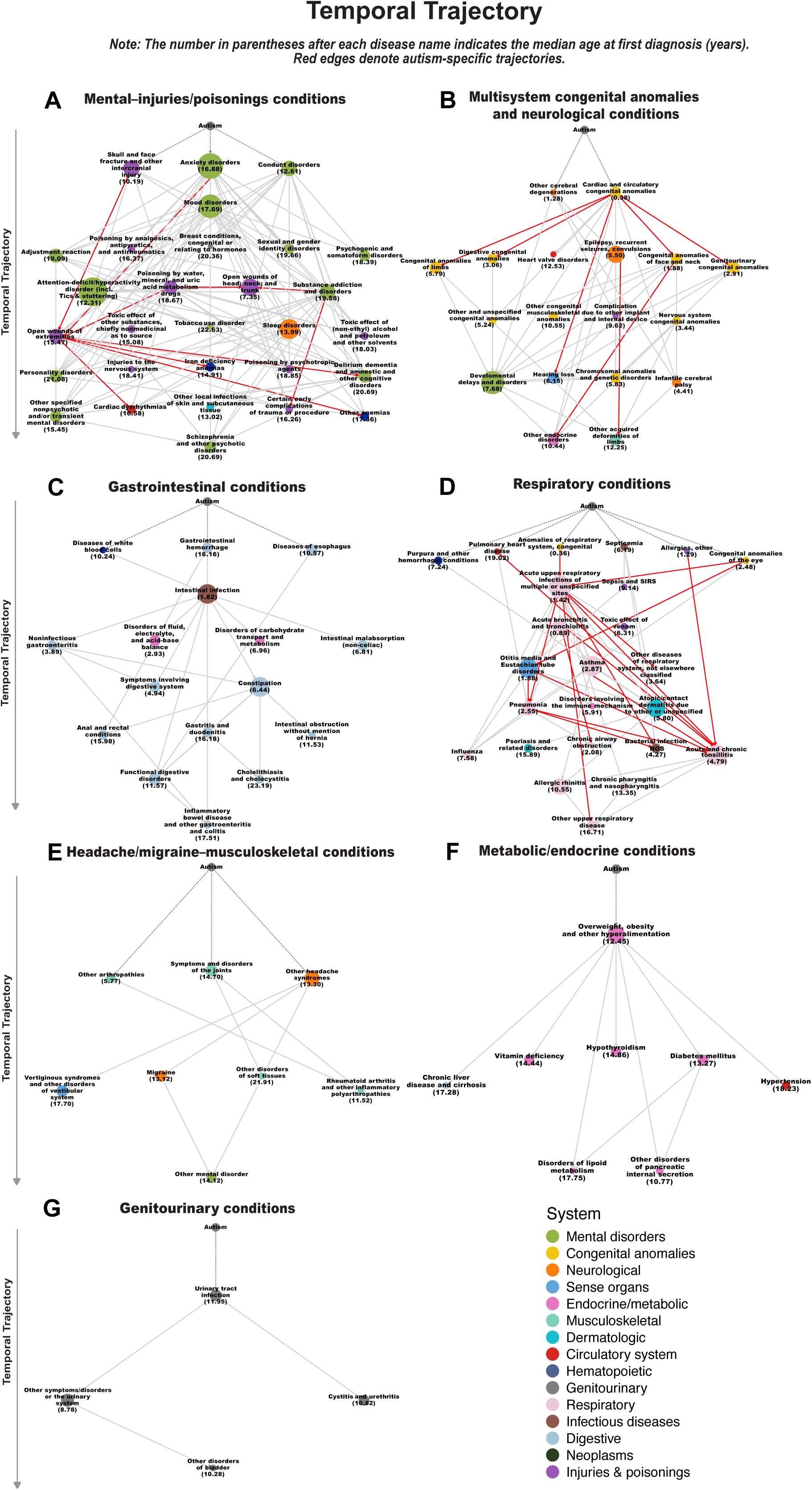
Disease trajectories among autistic individuals. (A–G) Temporal trajectories of medical conditions within seven comorbidity clusters. Each node represents a phecode-defined medical condition, and node color indicates disease system. Directed edges indicate significant temporal ordering between conditions, with red edges denoting autism-specific trajectories. The number in parentheses after each disease name indicates the median age at first diagnosis (years) of each disease.

To assess whether these temporal patterns were specific to autism, we tested interactions by autism status, quantifying whether directional associations were stronger among individuals with autism compared with matched peers. Within the seven clusters, we identified 34 relationships with significantly stronger directional associations in the autism group (all P for interaction <0.05 after BH correction & OR>1; red edges in Figure 4; Table S4).

The respiratory cluster exhibited the greatest number of directional associations that were stronger among individuals with autism, predominantly involving a broad spectrum of respiratory tract infections (Figure 4D). Additional autism-specific strengthening of directional associations was observed for malformations, including links between cardiac malformations and multiple congenital anomalies, as well as the co-occurrence of epilepsy and acquired limb deformities (Figure 4B). There was also evidence of stronger directional associations for a range of psychiatric and injury-related conditions (Figure 4A).

### Subgroup and sensitivity analyses

#### 1. Sex-stratified analyses

The overall cluster structure and temporal trajectories were largely consistent between males (N=66,606) and females (N=34,915) with autism, with disease system–level clusters reproducible across sexes (Figure S5; Table S5). Notable differences were nevertheless observed. The genitourinary cluster showed clear sex-specific composition, with females additionally encompassing reproductive conditions, including inflammatory diseases of the pelvic organs (Figure S5G). In males, a distinct dermatologic cluster emerged (Figure S5L), whereas females exhibited a more pronounced pain and inflammatory musculoskeletal profile (Figure S4E).

#### 2. Analyses stratified by co-occurring intellectual disability (ID)

The subgroup of individuals with autism without intellectual disability (N=85,031) largely recapitulated the main findings (Figure S6H–M; Table S6). In contrast, the subgroup with co-occurring intellectual disability (N=16,490) exhibited a markedly sparser cluster structure, with reduced cross-comorbidity correlations and less clearly defined temporal sequences of comorbidities (Figure S6A–G; Table S6). This pattern may reflect phenotypic differences in individuals with co-occurring intellectual disability, as well as the smaller sample size.

#### 3. Sensitivity analysis redefining the start of follow-up

In the sensitivity analysis in which follow-up was initiated at the date of autism diagnosis rather than birth, the PheWAS findings were broadly consistent with those from the main analysis (Figure S7; Table S7). The major disease-system patterns and the phenotypes most strongly associated with autism were largely retained, although some associations showed attenuated effect estimates and fewer phenotypes reached statistical significance. These results suggest that the overall comorbidity profile identified in the main analysis was not primarily driven by health records accumulated before autism diagnosis, and was generally robust to re-defining the start of follow-up.

## Discussion

In this nationwide population-based study, we mapped recorded morbidity and healthcare trajectories in more than 100,000 autistic individuals by integrating phenome-wide risk profiling, comorbidity network modelling, and temporally ordered diagnostic trajectories. Three findings emerged. First, autistic individuals had higher recorded risks for most examined conditions across organ systems (165 of 187 examined conditions), underscoring the breadth of healthcare-recorded morbidity in autism. Second, this excess morbidity was not randomly distributed across the phenome, but organized into seven clinically coherent clusters spanning psychiatric and injury-related conditions, congenital and neurological conditions, respiratory conditions, gastrointestinal conditions, genitourinary conditions, headache and musculoskeletal conditions, and metabolic and endocrine conditions. Third, the diagnostic trajectories enriched in autism were concentrated within a limited number of domains, suggesting that morbidity in autism may unfold through recurring healthcare pathways rather than through a diffuse accumulation of unrelated conditions.

### Comorbidity networks and trajectories

We identified seven comorbidity clusters, most of which were characterized by disease system-dominant modules, indicating a structured rather than diffuse pattern of co-occurrence(24). That is, instead of a high degree of comorbidity across disease-systems, we find that many clusters originate from single (e.g., cardiometabolic comorbidities) or dual disease-systems (e.g., psychiatric comorbidities and injuries). This is consistent with what has been seen in other psychiatric conditions, such as major depressive disorder (25), but also the general population (26). This may suggest that the experienced comorbidity in autism is often concentrated around a single condition or clinical domain and subsequently results in related complications surrounding the core comorbidity.

The observed separation into disease coherent clusters, alongside substantial between-cluster differences in the median age at first recorded diagnosis, suggests that clustering captures not only comorbidity structure but also cluster-specific windows of clinical presentation (27). While diagnostic timing can be influenced by ascertainment and healthcare use, the resulting temporal profiles offer a pragmatic scaffold for stratified monitoring and hypothesis generation regarding developmentally patterned comorbidity in autism (28). The observation that developmental delays and epilepsy were strongly connected to a broad spectrum of congenital anomalies—and clustered with multisystem congenital anomalies rather than with psychiatric/injury-related conditions—may reflect an underlying neurodevelopmental–genetic architecture shared across these phenotypes. This interpretation is consistent with prior evidence that rare pathogenic variants, particularly copy-number variants (CNVs) and other genomic alterations, contribute to a spectrum spanning congenital anomalies, developmental delay/intellectual disability, autism-related traits, and epilepsy, supporting the notion that these co-occurring conditions can arise from overlapping developmental and genetic mechanisms (29–35).

Consistent with previous studies, psychiatric conditions were among the most prevalent comorbidities in autistic individuals. This pattern probably reflects a combination of shared liability, heightened clinical surveillance, and increased vulnerability to mood and anxiety-related conditions arising from stigma, trauma, social exclusion, and other challenges experienced by autistic individuals (36–39). One of the major comorbidity clusters identified in our study comprised psychiatric disorders and external causes of morbidity, including injuries and poisonings. The close clustering and similar diagnostic timing of these conditions may reflect several, non-mutually exclusive processes: psychiatric comorbidity may increase injury risk through impulsivity, inattention, or substance use; injury-related healthcare encounters may facilitate detection of mental and behavioural conditions; and both may arise from shared social and environmental risk factors (40–42). This cluster underscores the need for proactive risk-mitigation strategies tailored to autistic populations, particularly among individuals with co-occurring psychiatric conditions (43).

Nutritional deficiencies also appeared within this cluster. Our data cannot determine whether psychiatric comorbidity contributes to later nutritional deficiencies through eating difficulties or altered self-care, whether nutritional deficiencies increase vulnerability to psychiatric morbidity, or whether they serve as early markers of broader comorbidity accumulation in autistic individuals (44–49). For example, the clustering of iron deficiency with mental and behavioural conditions, injuries, and poisonings may point to behavioural pathways, such as sensory-driven food selectivity, underlying nutritional deficits in autistic individuals. More broadly, these findings highlight the importance of monitoring nutritional status in autistic individuals, particularly in the presence of psychiatric comorbidity.

Another major cluster was characterized predominantly by gastrointestinal conditions. Intestinal infection, functional digestive disorders, and constipation showed the three highest centrality measures within this cluster. Notably, constipation is among the most commonly reported gastrointestinal symptoms in autistic children (50–52). Its frequent occurrence may partly reflect autism-related feeding and behavioral characteristics, including food selectivity, restricted dietary variety, sensory sensitivities affecting food acceptance, low fiber or fluid intake, and toileting-related difficulties (50, 53–55).

The temporal structure of these clusters may help contextualize how comorbidities accumulate across development in autistic children and young people. We identified 34 diagnostic trajectories that were more prominent among autistic individuals than controls. Consistent with previous work (27, 56), these autism-enriched trajectories were not diffusely distributed across the disease spectrum, but concentrated within a limited number of clinical domains. The strongest enrichment was observed for trajectories involving injuries or poisonings and respiratory tract conditions, suggesting that injury-related encounters and recurrent respiratory morbidity represent important points for clinical monitoring in autistic individuals (57–61). Associations were also stronger for autistic individuals within the congenital anomaly domain, which may reflect an underlying neurodevelopmental–genetic architecture that links multisystem malformations with later neurological and developmental manifestations(62–64). Collectively, these findings indicate that excess morbidity in autism may cluster within a small set of key domains rather than being uniformly distributed across organ systems.

Subgroup and sensitivity analyses indicated broadly similar morbidity structures, with some group-specific findings. Across strata defined by sex and co-occurring ID, the major network modules and temporal trajectory domains were generally reproducible, although smaller subgroups, particularly individuals with co-occurring ID, showed sparser network structures. Several subgroup-specific patterns were observed, including sex-related differences in genito-urinary, dermatologic, and musculoskeletal-inflammatory profiles. Overall, these subgroup findings may reflect genuine phenotypic heterogeneity, but they could also be influenced by sample size and follow-up-related factors. The sensitivity analysis using autism diagnosis as the start of follow-up produced broadly consistent PheWAS findings, supporting the robustness of the main morbidity profile.

### Strengths and limitations

This study has several strengths. It leveraged nationwide register data with extended followup, enabling population-level characterization of diagnosed disease burden among autistic individuals. By integrating phenome-wide risk profiling, comorbidity clustering, and temporal trajectory analysis, the study provides a broader view of morbidity in autism than previous condition-by-condition investigations. This approach allowed us to identify not only individual diagnoses associated with autism, but also broader patterns of co-occurring and temporally linked conditions across development.

However, several limitations should also be considered. First, the analyses relied on diagnoses recorded in specialist outpatient and inpatient care. This may underestimate conditions managed primarily in primary care, such as type 2 diabetes, and may miss milder but prevalent conditions that do not require specialist care yet still contribute substantially to everyday disease burden. Future studies incorporating primary care data will therefore provide an important complement to the patterns reported here. Second, the results capture diagnosed morbidity rather than the full underlying burden of symptoms. Swedish registers lack detailed information on symptom severity, functional impairment, lifestyle factors, and patient-reported outcomes, which limits interpretation of the clinical impact of some comorbidity patterns. Finally, we did not examine interventions, medication use, or care pathways. These data are available in Swedish registers but would require substantial harmonization to be incorporated into a coherent phenome-wide framework. Future work linking diagnoses with treatments, medication exposure, and healthcare utilization could provide an even richer picture of the clinical trajectories experienced by autistic individuals.

An important consideration is that the observed comorbidity patterns and temporal trajectories reflect recorded healthcare encounters rather than necessarily the underlying order of disease onset or progression. Differences in healthcare access, referral patterns, diagnostic intensity, and coding practices are likely to shape the observed comorbidity landscape. Autistic individuals may have more frequent contact with specialist services, greater clinical surveillance, and different thresholds for diagnostic coding than matched peers. Conversely, communication differences, diagnostic overshadowing, and barriers to care may delay or obscure recognition of some conditions. The resulting patterns therefore reflect the intersection of underlying morbidity and healthcare-system response. This does not diminish their clinical importance, as recorded diagnoses form the basis for treatment, follow-up, and resource allocation. However, elevated risks and temporal sequences should not be interpreted as direct estimates of biological disease incidence or disease progression without considering ascertainment and care-seeking processes. Whether some conditions should have been detected earlier, or whether specific sequences indicate missed opportunities for earlier intervention, for example, comorbid ADHD being diagnosed only after several other psychiatric contacts, cannot be determined from the observed comorbidity patterns alone. Nevertheless, these trajectories provide an empirical basis for identifying recurring diagnostic pathways and for informing future work on earlier recognition, monitoring, and care coordination in autistic individuals.

An additional interpretive consideration is that autism is itself clinically ascertained over development. Some recorded comorbidities may therefore precede the autism diagnosis and contribute to clinical recognition, referral, or diagnostic evaluation, rather than representing consequences of autism. This is particularly relevant for neurodevelopmental, psychiatric, congenital, and neurological conditions that may increase contact with specialist services. Accordingly, the observed trajectories should be interpreted as diagnostic and healthcare pathways among individuals who were ultimately diagnosed with autism, not as evidence that autism causes subsequent morbidity. This distinction is central when using register-based data to study developmental conditions whose diagnosis may occur after several years of healthcare contact.

## Conclusion

This study provides a population-level characterization of multimorbidity in autism, showing that excess morbidity concentrates within clinically coherent clusters and recurrent diagnostic trajectories rather than being uniformly distributed across organ systems. Although these patterns reflect recorded care and should not be interpreted as disease progression, they identify clinically salient pathways that may inform future work on stage-specific monitoring, care co-ordination, and mechanisms underlying excess morbidity in autistic individuals.

## Methods

### Data sources

The Swedish National Patient Register contains nationwide health records with nearly complete coverage of inpatient care since 1987 and outpatient specialist care since 2001. From this register, we identified all autism diagnoses and all other diagnostic codes used in this study, together with their recorded dates. Vital status and dates of emigration were obtained from the Total Population Register (established in 1968). Sociodemographic data were retrieved from the Longitudinal Integrated Database for Health Insurance and Labour Market Studies (LISA, available since 1990), including maternal and paternal income and parental highest attained education level. For education, we defined parental education as the highest educational level attained by either parent between 1990 and 2020, coded as an ordinal variable from 1 to 7 representing increasing educational attainment: 1 = not completed compulsory education (< 9 years), 2 = completed compulsory education (9 years), 3 = upper secondary (2 years), 4 = upper secondary (3 years), 5 = college/university < 3 years, 6 = college/university ≥ 3 years, 7 = research education. For income, we obtained annual disposable income for each year from 1990 to 2020 and ranked it between 0 (lowest) and 1 (highest) relative to the entire Swedish population in that specific year, thereby accounting for outliers and inflation. To minimize missingness, we assigned family income by anchoring to the child’s birth year and selecting the income record from the year closest to the date of birth with available income data for at least one parent (ties were resolved by choosing the later year); if both parents had data in that year, we used the higher ranked value, and if only one parent had data, that value was used. Family income was then defined as the quintile corresponding to this selected ranked measure and assigned to the child. All registers are mandated by law and are virtually complete. Missing sociodemographic data primarily occurs among recent immigrants, individuals with unrecorded paternal information, or parents working abroad while residing in Sweden (65). In the analyses, missing values were treated as a separate category. The Swedish Ethical Review Authority (2020-05516 & 2022-05648-02) approved the study and waived the need for informed consent.

### Participants and follow-up

We included all individuals born in Sweden between 1987 and 2020 who were diagnosed with autism in specialized outpatient or inpatient care any time through 31 December 2021 (N = 101,521). Diagnoses of autism were ascertained using the Swedish revisions of the International Classification of Diseases (ICD-9 code 299; ICD-10 code F84). For each individual with autism, we randomly selected ten unaffected individuals from the Total Population Register, individually matched on sex and birth year and month (N = 958, 093). Except for in sensitivity analysis, we began follow up at birth rather than at diagnosis date since autism is generally assumed to be present from birth, although often only recognized later(66, 67). Matched affected individuals were required to have no record of an autism diagnosis at any point during the study period and to be alive and not emigrated from Sweden at the diagnosis date of their matched autism case. All participants were followed for medical conditions from birth until death, emigration, or the end of follow-up (31 December 2021), whichever occurred first.

Medical conditions during follow-up were identified through the National Patient Register, classified according to three-digit ICD-9 (001–999) and ICD-10 (A00–N99) codes. All ICD codes were mapped to their corresponding phecodes (Table S8)(21), with the categories of pregnancy complications, symptoms and others excluded (Table S9). To avoid mixing sex-specific conditions in the same framework, we removed sex-specific conditions in the main analysis and most of the subgroup analyses, but included them in the sex-stratified subgroup analyses. A total of 16,795 ICD-9 diagnostic occurrences (8.43%) and 688,579 ICD-10 diagnostic occurrences (5.44%) remained unmapped to phecodes (Table S10). To protect individual-level privacy, the Swedish National Board of Health and Welfare truncated some rare diagnostic codes (e.g., M77–M79). Because the exact underlying diagnoses could not be determined for these truncated codes, we did not include them in the analysis to ensure greater precision and avoid misclassification.

### Disease network analysis

We implemented a three-step analytical pipeline: (i) a phenome-wide association study (PheWAS), (ii) disease network analysis and (iii) disease trajectory analysis. Detailed descriptions of these approaches have been outlined elsewhere(68). We further assessed effect modification by autism status in the disease trajectory analysis to identify autism-specific temporal trajectories. Across all analyses, multiplicity was controlled using the Benjamini–Hochberg false discovery rate (FDR) at 5%, and q values are reported throughout.

#### 1. Phenome-wide association study

We began by performing a PheWAS to identify conditions more common among individuals with autism compared with the matched unaffected individuals. To ensure adequate power, only conditions with incidence ≥0.1% among individuals with autism were retained. Associations were estimated using Cox proportional hazards models with attained age as the time scale, stratified by matching set (sex and birth month and year) and adjusted for birth country, highest parental education, and family income. Conditions advancing to downstream analyses were those showing statistically significant positive associations with autism after multiple-testing correction (HR > 1 & q < 0.05).

Among retained conditions, all possible disease pairs - denoted disease 1 (D1) and disease 2 (D2) - were constructed, considering only pairs with co-occurrence incidence ≥0.1% among individuals with autism. Comorbidity strength was quantified using relative risk (RR) and Pearson’s φ-correlation within individuals with autism. Pairs retained for network and trajectory analyses satisfied RR > 1, φ > 0, and q < 0.05. In other words, we excluded disease pairs that failed a simple univariate test, because they likely reflect stochastic events or weak links that would not hold up in later comorbidity network and disease trajectory analyses, and add little to overall comorbidity patterns.

#### 2. Comorbidity Network Analysis

Non-temporal associations were estimated within individuals with autism using unconditional logistic regression with partial correlation approach(69). To account for confounding arising from the co-occurrence of other conditions, the first 20 principal components derived from all retained medical conditions in the network were included as covariates. In addition, the models were adjusted for birth country, highest parental education, family income, age at diagnosis, sex, and birth year (categorized into seven groups). Pairs with positive associations (OR>1) and q < 0.05 were assembled into an undirected comorbidity network. Network modules - communities of conditions with high within-module connectivity - were identified using the Louvain algorithm(70), revealing clusters of frequently co-occurring conditions in individuals with autism.

To identify hub conditions in the comorbidity network, we quantified node centrality using node strength, defined as the sum of incident edge weights for each node, where edge weights represented comorbidity strength between paired conditions.

#### 3. Temporal Trajectory Analysis

To assess temporal directionality, binomial tests were applied to each qualifying disease pair, defined using a time window of 30 days to 5 years, to determine whether D2 more often followed D1 than vice versa among individuals with autism. Pairs with a consistent temporal order (D1 → D2) were analyzed using nested case–control design, treating D2 as the outcome and D1 as the exposure. For each D2 case, up to five controls were selected by incidence density sampling, matched on sex and birth year. Conditional logistic regression models were adjusted for birth country, highest parental education, family income, age at autism diagnosis, and the first 20 principal components derived from all other retained medical conditions under the partial correlation framework. Directed links satisfying OR > 1 and q < 0.05 were concatenated into multi-step disease trajectories (e.g., D1 → D2 → D3), providing a high-resolution map of sequential diagnoses in autism. For each condition in the trajectory, we calculated the median age at first diagnosis (years), defined as the time from birth to the first recorded diagnosis among individuals diagnosed during follow-up.

#### 4. Autism-specific trajectories

To identify autism-specific temporal transition patterns, we reincluded individuals with autism and the reference group and tested whether the disease A → disease B association differed by autism status using an interaction model. Candidate directed disease pairs (disease A → disease B) were restricted to those identified as significant in the main trajectory analysis. For each disease pair, we fit a logistic regression model on an outcome condition (e.g., disease B) with the predictors being autism status, an indicator for exposure (e.g., disease A), and their interaction (e.g., autism status × disease A), with adjustment for birth country, highest parental education, family income, sex, birth year group. An association was classified as autism-specific if the interaction term was positive and statistically significant (P for interaction <0.05 after B-H correction).

### Subgroup and sensitivity analyses

Subgroup analyses were conducted by stratifying individuals with autism according to (1) sex (male/female), (2) co-occurring ID (yes/no). Within each subgroup, we repeated the PheWAS analyses, comorbidity network and trajectory analyses using the same analytic procedures as in the main analysis. In a sensitivity analysis, follow-up was initiated at the date of autism diagnosis rather than birth, and the main analysis were repeated.

## Data Availability

The individual-level data used in this study were obtained from Swedish national registers and are not publicly available due to legal and ethical restrictions. Access to these data is subject to approval by the relevant Swedish authorities. Researchers interested in accessing the data may apply directly to the respective data-holding authorities in accordance with applicable regulations.

## Statements and Declarations

### 7. Ethics statement

All research was performed in accordance with relevant guidelines/regulations. The applied analysis was approved by the Regional Ethical Review Board in Stockholm (2020-05516 & 2022-05648-02).

### 8. Author Contributions

LKH, TR, FL and VHA designed the study. LKH and VHA conducted data analysis, with ZZX contributing to part of the data analysis. LKH, TR and VHA drafted the manuscript. LKH, TR, CH, FF, JG, ZZX, YWP, DS, BKL, FL and VHA contributed to the interpretation of the data and critically and substantively revised the manuscript. FL and VHA contributed to the work as senior investigators. They designed the study, supervised the data analyses, and supervised the drafting and revising of the manuscript. The corresponding author attests that all the listed authors meet authorship criteria and that no others meeting the criteria have been omitted. All authors read and approved the final manuscript.

### 9. Availability of data and material

The data used for the applied example are publicly unavailable according to regulations under Swedish law. Readers interested in obtaining microdata may seek similar approvals and inquire through Statistics Sweden.

