## Supplemental Information for "High-Resolution Mapping of Healthcare Pathways in Autism"

* Shared first authors

\# Shared senior authors

† Corresponding authors

**Supplementary Figures**

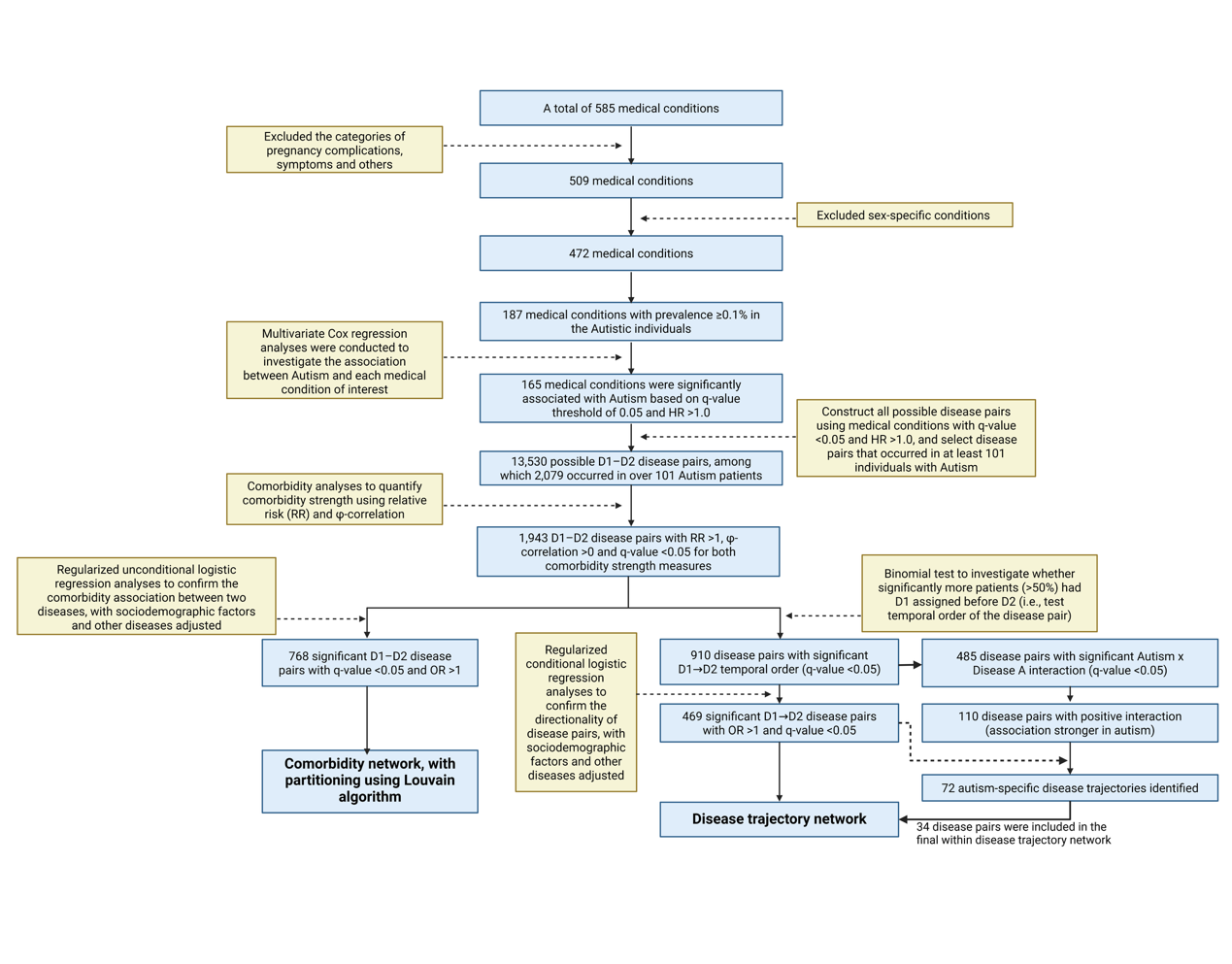

**Supplementary Figure 1** Flowchart for three-dimensional disease network analysis

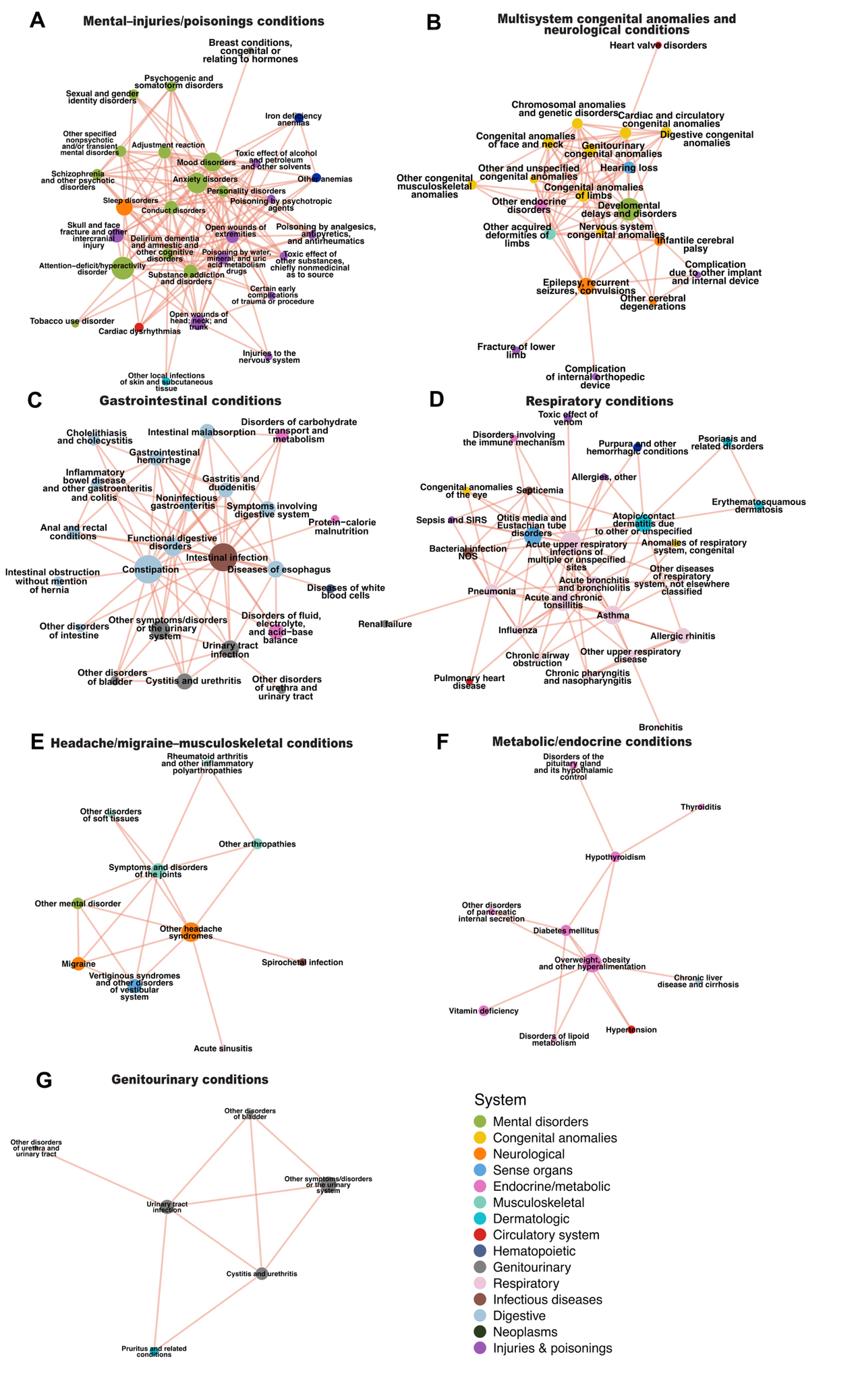

**Supplementary Figure 2** Within-cluster comorbidity networks among autistic individuals

Each node represents a disease; node size reflects disease prevalence and node color denotes the disease category. The network was partitioned into seven clusters using the Louvain algorithm; the diseases contained within each cluster are labeled on the nodes.

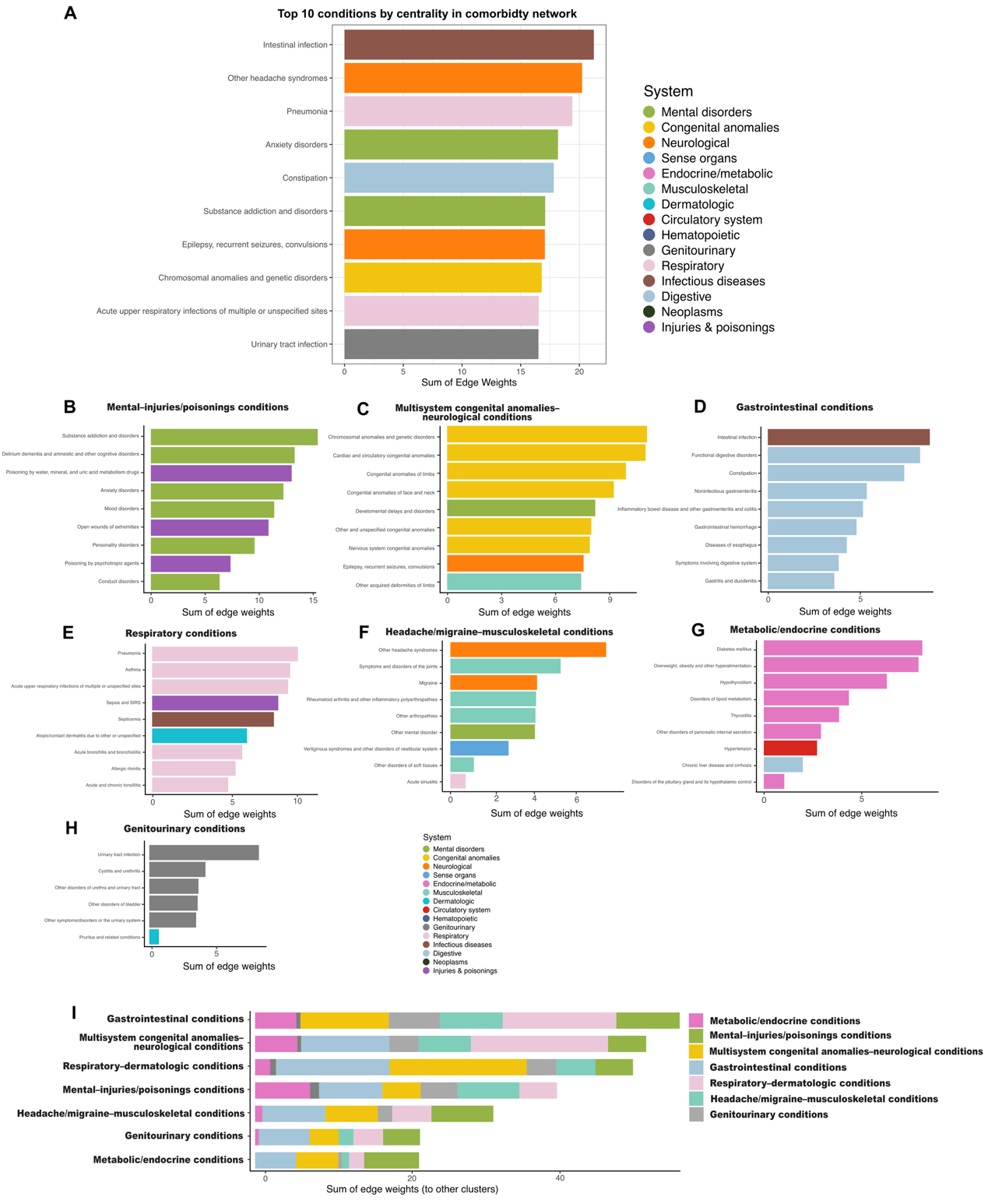

**Supplementary Figure 3** Medical conditions with the highest centrality within clusters and connectivity between clusters.

(A) Top 10 medical conditions with the highest node centrality in the overall comorbidity network among autistic individuals. Centrality was quantified as node strength, defined as the sum of edge weights connected to each node, where edge weights represent the strength of comorbidity between paired conditions.

(B–G) Medical conditions with the highest centrality within each disease cluster: mental–injuries/poisonings conditions (B), multisystem congenital anomalies–neurological conditions (C), gastrointestinal conditions (D), respiratory conditions (E), headache/migraine–musculoskeletal conditions (F), metabolic/endocrine conditions (G), and genitourinary conditions (H).

(H) Between-cluster connectivity showing the summed edge weights linking conditions in each cluster to conditions in other clusters.

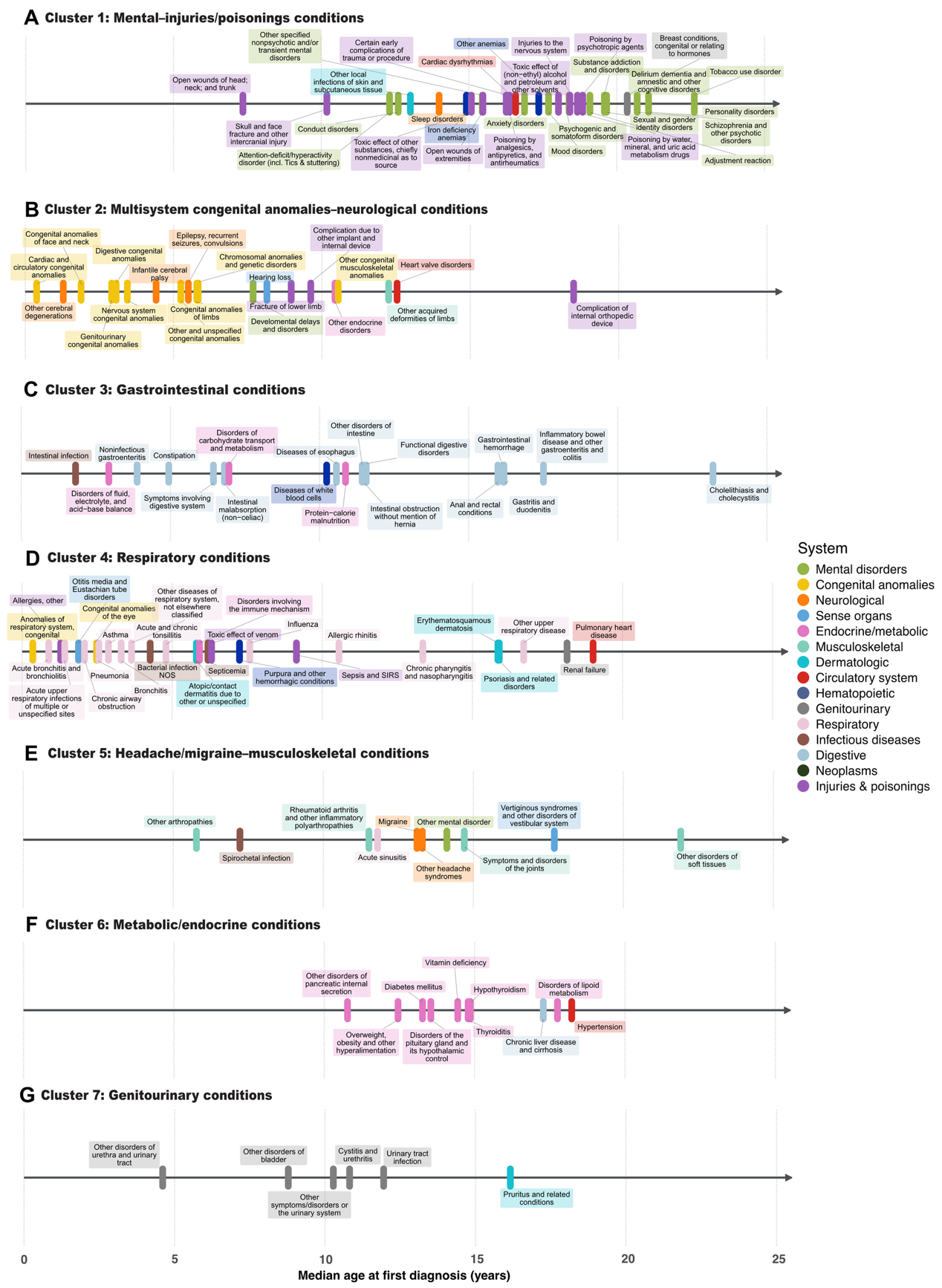

**Supplementary Figure 4** Median age at first diagnosis of medical conditions across clusters in autistic individuals

This figure shows the median age at first diagnosis of individual medical conditions within each comorbidity cluster identified among autistic individuals. Each panel represents one cluster: (A) mental–injuries/poisonings conditions, (B) multisystem congenital anomalies–neurological conditions, (C) gastrointestinal conditions, (D) respiratory conditions, (E) headache/migraine–musculoskeletal conditions, and (F) metabolic/endocrine conditions, and (G) genitourinary conditions. Each colored label represents one phecode-defined medical condition, positioned along the x axis according to its median age at first diagnosis in years. Label colors indicate disease system categories. The horizontal axis represents median age at first diagnosis (years).

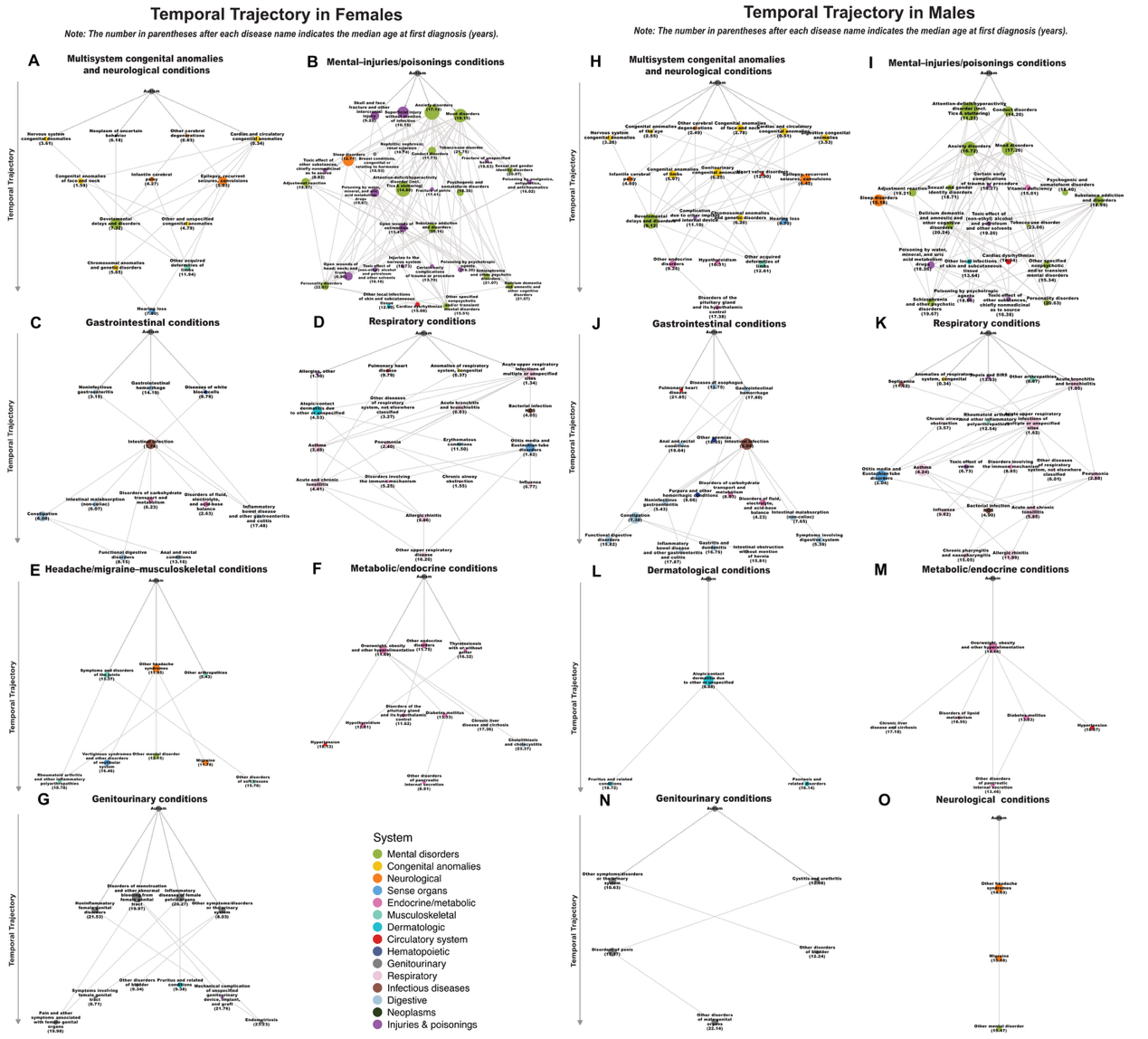

**Supplementary Figure 5** Disease trajectories among autistic individuals stratified by sex.

Temporal disease trajectories identified among autistic individuals in females (A–G) and males (H–O). Each node represents a disease, with node color indicating disease category. The number in parentheses after each disease name indicates the median age at first diagnosis (years).

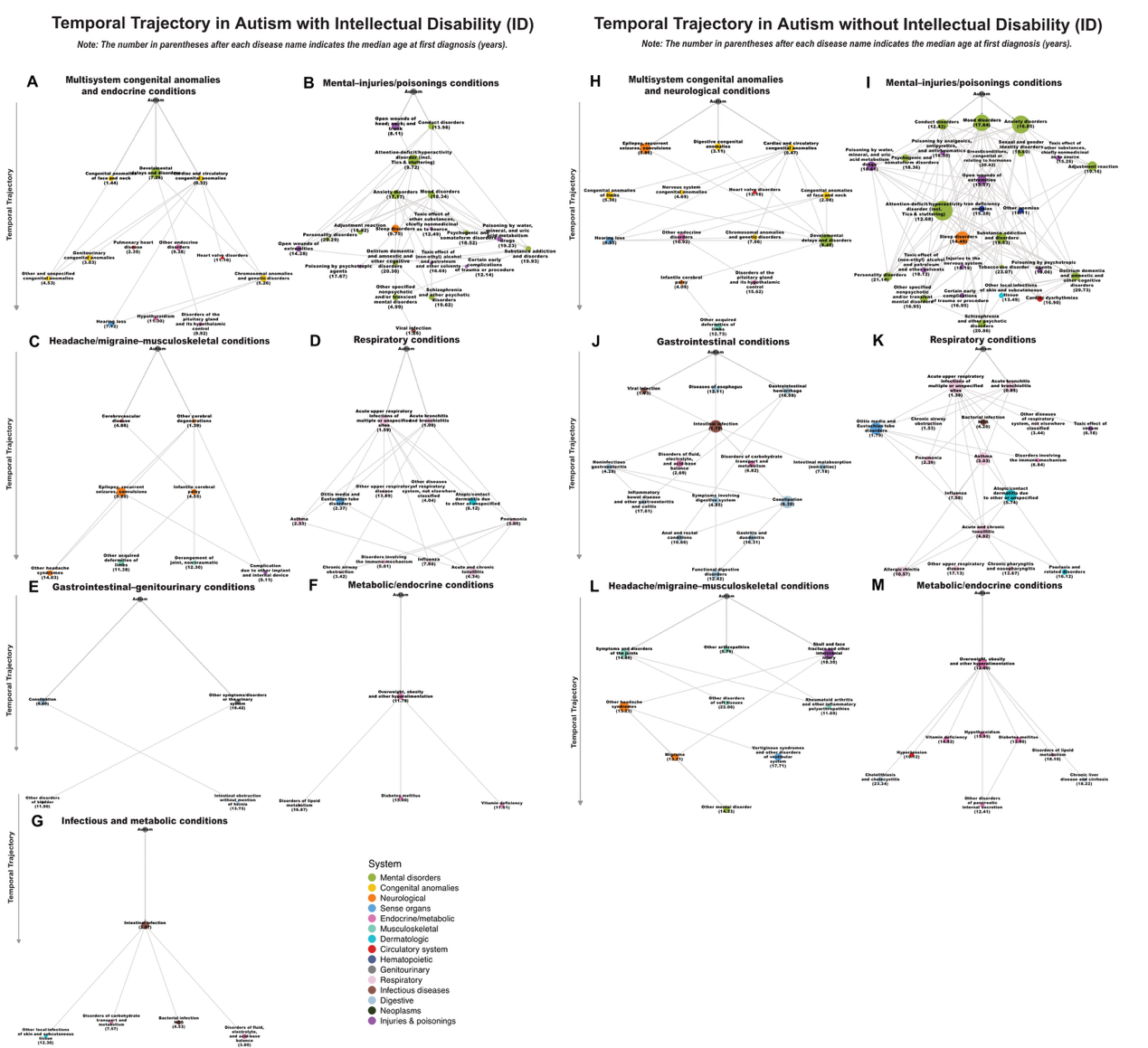

**Supplementary Figure 6** Disease trajectories among autistic individuals stratified by co-occurring intellectual disability status.

Temporal disease trajectories identified among autistic individuals with intellectual disability (A–G) and without intellectual disability (H–M). Each node represents a disease, with node color indicating disease category. The number in parentheses after each disease name indicates the median age at first diagnosis (years).

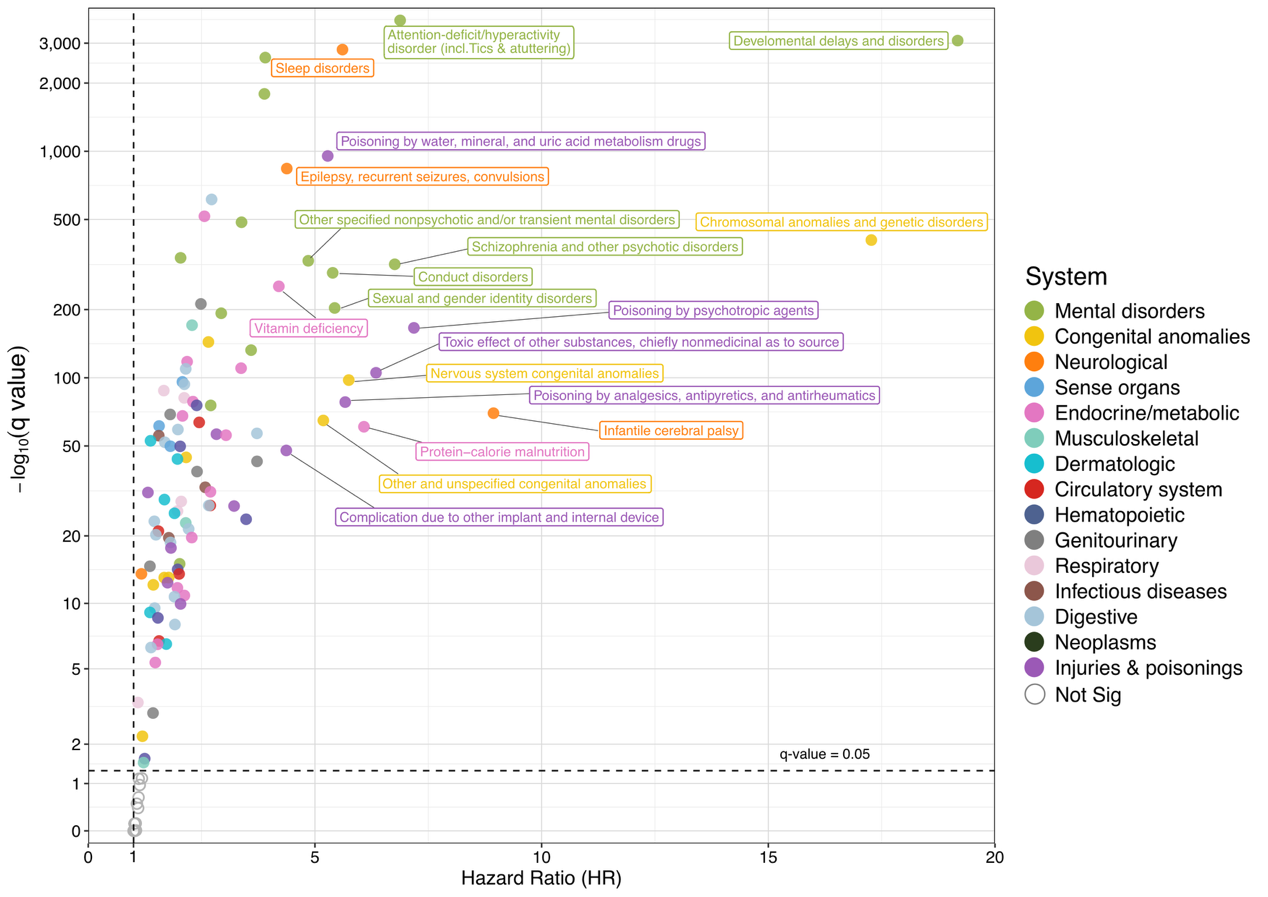

**Supplementary Figure 7** Sensitivity analysis of PheWAS results using autism diagnosis as the start of follow-up

Scatter plot of hazard ratios (HRs) and corresponding significance levels for 148 phecode-defined medical conditions in the sensitivity analysis, in which follow-up was initiated at the date of autism diagnosis rather than birth. Each point represents one phecode-defined condition, with the x-axis indicating the HR and the y-axis showing the corresponding −log10(q value). Colors denote disease systems. The vertical dashed line indicates HR = 1, and the horizontal dashed line indicates the false discovery rate threshold of q = 0.05. Selected conditions with the strongest associations are labeled.

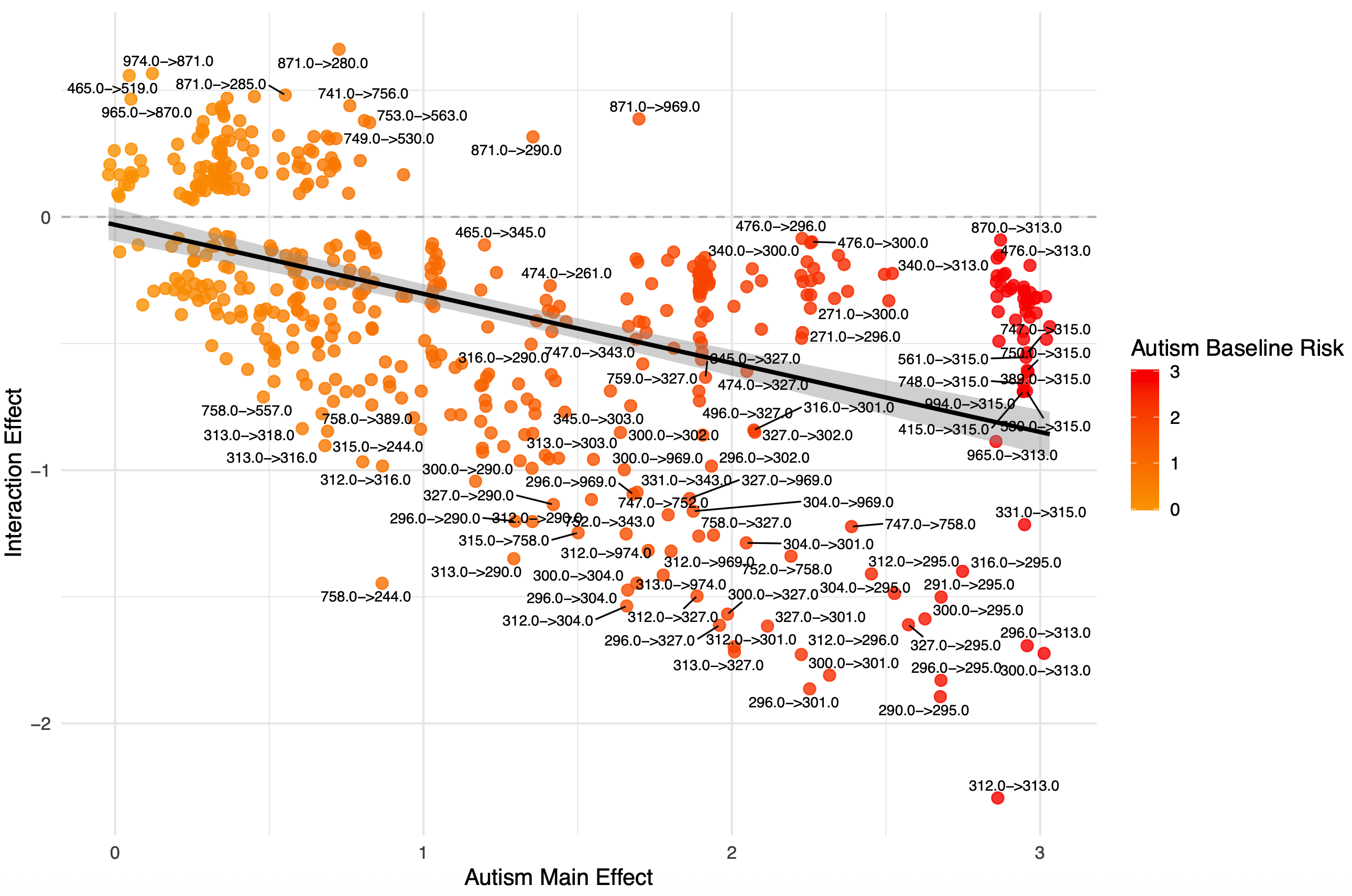

**Supplementary Figure 8** Relationship between autism main effects and autism-specific interaction effects across disease pairs.

Each point represents a directed disease pair (A→B) identified in the trajectory analysis. The x-axis shows the autism main effect (β1), representing the overall association between autism status and the subsequent disease. The y-axis shows the autism-specific interaction effect (β3) from the interaction model (autism status × disease A), indicating whether the temporal association between diseases differs between autistic individuals and the reference group. Point color represents the baseline risk of the disease among autistic individuals. The solid line indicates the fitted linear regression, and the shaded area represents the 95% confidence interval. The solid line represents the fitted linear regression, and the shaded area indicates the 95% confidence interval. Higher autism main effects were associated with more negative autism-specific interaction effects (β = −0.273, SE = 0.022, P < 2×10⁻¹⁶; R² = 0.239).
